# Stochastic challenges to interrupting helminth transmission

**DOI:** 10.1101/2019.12.17.19013490

**Authors:** Robert J. Hardwick, Marleen Werkman, James E. Truscott, Roy M. Anderson

## Abstract

Predicting the effect of different programmes designed to control both the morbidity induced by helminth infections and parasite transmission is greatly facilitated by the use of mathematical models of transmission and control impact. In such models, it is essential to account for the many sources of uncertainty — natural, or otherwise — to ensure robustness in prediction and to accurately depict variation around an expected outcome. In this paper, we investigate how well the standard deterministic models match the predictions made using individual-based stochastic simulations. We also explore how well concepts which derive from deterministic models, such as ‘breakpoints’ in transmission, apply in the stochastic world. Employing an individual-based stochastic model framework we also investigate how transmission and control are affected by the migration of infected people into a defined community. To give our study focus we consider the control of soil-transmitted helminths (STH) by mass drug administration (MDA), though our methodology is readily applicable to the other helminth species such as the schistosome parasites and the filarial worms. We show it is possible to theoretically define a ‘stochastic breakpoint’ where much noise surrounds the expected deterministic breakpoint. We also discuss the concept of the ‘interruption of transmission’ independent of the ‘breakpoint’ concept where analyses of model behaviour illustrate the current limitations of deterministic models to account for the ‘fade-out’ or transmission extinction behaviour in simulations. Our analysis of migration confirms a relationship between the critical infected human migration rate scale (i.e., order of magnitude) per unit of time and the death rate of infective stages that are released into the free-living environment. This relationship is shown to determine the likelihood that control activities aim at chemotherapeutic treatment of the human host will eliminate transmission. The development of a new stochastic simulation code for STH in the form of a publicly-available open-source python package which includes features to incorporate many population stratifications, different control interventions including mass drug administration (with defined frequency, coverage levels and compliance patterns) and inter-village human migration is also described.

## 1. Introduction

Helminthiases are a class of the neglected tropical diseases (NTDs) that affect billions of humans and animals worldwide. One group of worms, the soil-transmitted helminths (STH), are especially prevalent and are transmitted through the ingestion of eggs (for *Ascaris lumbricoides* or *Trichuris trichuria*) or larvae (in the case of the hookworms: *Necator americanus* and *Ancylostoma duodenale*). They are estimated to affect 1.45 billion people at present [1, 2].

In recent years, mathematical models of both infectious disease transmission [3, 4, 5] and intervention impact have been widely used in infectious disease epidemiological studies and public health policy formulation [6, 7, 8, 9, 10]. Their use in the study of the NTDs is more recent, where the variety of approaches implemented have included both deterministic [11, 7] and individual-based stochastic simulation models [12, 13, 14,15, 16, 17, 18]. Much progress in model formulation, parameter estimation and application has been made over the last 20 years. Model-based analyses are increasingly serving an important role in policy formulation and the evaluation of different control policies [19, 20, 21] as is well illustrated by the activities of the Bill and Melinda Gates Foundation funded NTD Modelling consortium [22]. Furthermore, following the London Declaration in 2010, which stimulated the expansion of large mass drug administration (MDA) programmes under the direction of World Health Organization (WHO) guidelines on treatment strategies [20, 23, 24], mathematical models have played an increasing role in determining how best to design and evaluate MDA programmes [25, 26, 27, 28, 29, 30, 31].

Two key concepts emerge from analyses of deterministic models. The first is the existence of a ‘breakpoint’ in transmission created by the dioecious nature of the worms (a male and female must be present in the same host to ensure the production of fertile eggs and pass to the external environment and form the pool of infectious material) which creates an unstable equilibrium separating the stable endemic infection equilibrium from the other stable equilibrium of parasite extinction. Independent of this breakpoint, a further epidemiological situation arises where only one stable equilibrium exists; namely, parasite and transmission extinction where the rate of infection is always too low to sustain the parasites in the human host. In this case transmission is interrupted, since an adult female worm in the human host on average, produces too few offspring to ensure one of her offspring matures in the human host to perpetuate the lifecycle. For many viral and bacterial infections (the microparasites), this is the situation where the basic reproduction number, *R*_0_ < 1, where *R*_0_ describes the generation of secondary cases. For the macroparasitic helminths, the concept of *R*_0_ needs modification since it describes the average number of female offspring, produced by female worms, that survive to reproductive maturity by infecting a new human host and maturing within it [7].

The concept of *R*_0_ is further modified by two density dependent processes: acting on fecundity as a population regulatory factor and acting via the dioecious nature of the worm. Because of the action of these two processes, the critical *R*_0_ value to sustain transmission is greater than unity in value. These two concepts are illustrated diagrammatically in Fig. 1. Note, in particular, how the darker red shaded region — which demarcates the *R*_0_ < 1 region — is replaced with a lighter shaded red ‘breakpoint’ region once these density dependent processes have been introduced.

**Figure 1:**
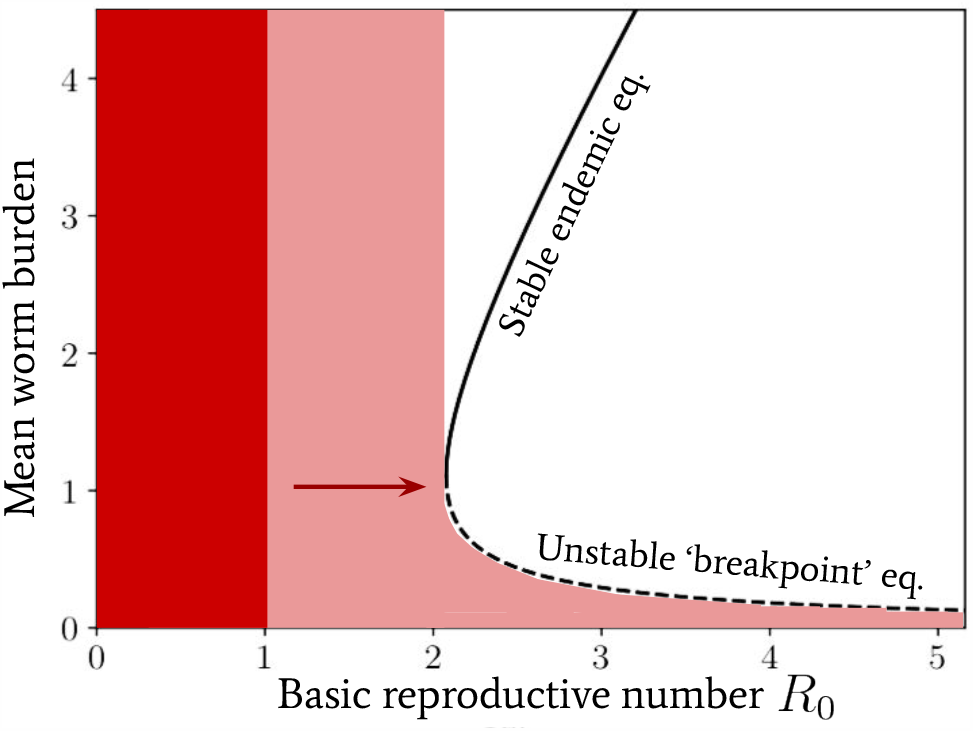
Diagram illustrating the stable endemic equilibrium (solid black line), unstable ‘breakpoint’ equilibrium (dashed black line) and insufficient *R*_0_ breakpoint. For reference, the transmission model illustrated is that of soil-transmitted helminth infections with the density dependent fecundity parameter γ = 0.08 and worm aggregation within hosts parameter *k* = 0.3.

The importance of these notions when we move from the deterministic world to a stochastic one is linked to explaining what the interruption of transmission really appears as in a noisy world full of variation and chance events. Even when the deterministic breakpoint is crossed, when its value is low due to high degrees of parasite aggregation in the human host (see Ref. [7]), stochastic noise may induce fluctuations where bounce back or extinction occur [30, 32]. Similarly, even when transmission is interrupted when the average female worm produces too few offspring to ensure the continuation of the life cycle, stochastic noise may not always result in parasite extinction in a defined location. In this paper, we shall analyze these situations with the use of a full individual-based stochastic model and certain approximations to the fully-simulated outcomes. In particular what is novel in our approach, is both looking at noise around the breakpoint and the transmission interruption state, and assessing how the migration of infected humans influences the likelihood of achieving effective control or even transmission interruption by MDA.

At present the demand is for increasing complexity in models to describe all known biological and epidemiological complexities including, for example: differing patterns of compliance to treatment; infected human migration patterns in and out of defined control activity regions; and various heterogeneities in human behaviour that influence transmission. In such circumstances, the temptation is to move to ever more complex simulation models, with a concomitant growth in parameters and the associated problems in measurement and estimation. In this paper we adopt a somewhat different approach, seeking to address the following fundamental questions:

- How well can deterministic models match the predictions made using stochastic simulations?
- Do the concepts such as ‘breakpoints’ in transmission still apply when stochasticity is introduced?
- How are transmission and control affected by infected human migration in a world of stochasticity?

In addressing these questions here we seek to ascertain the uncertainties that have the greatest impact on forecasts of NTD control initiatives both to enhance the quality of predictions and focus attention on what needs to be measured to improve accuracy.

The results described in this paper are underpinned by various analytical and numerical methods. We focus attention on stochastic models for STH infection and control, with epidemiological parameters set to those of the two hookworm species (*Necator americanus* and *Ancylostoma duodenale*). However, the methodologies developed are generalizable to other human helminth infections.

In Sec. 2 we introduce the mathematical model and formalism within which the first two main questions posed above are answered. In Sec. 3 we then extend this formalism to include a model for MDA control and infected human migration between reservoirs of infection, which we then use to answer the third question above. Lastly, in Sec. 4 we conclude with a summary of our findings and a discussion of future work.

## 2. A stochastic individual-based model

Following past publications [12, 26], the STH transmission model we introduce here is a stochastic individual-based analogue to the deterministic original given in Refs. [33, 11, 7].

For a recent review of stochastic STH models and their predictions concerning transmission dynamics and control impact, see Ref. [26]. The design of this model follows a similar pattern to Ref. [26], however, with some modifications made to the age structuring (by binning instead of a continuous function) and the novel addition of migration patterns between communities (which we introduce in more detail later).

Consider an ensemble of Poisson walkers, each representing individual worm hosts within a community, and each assigned a worm uptake rate λ_*jn*_Λ _*j*_(*t*) (or the ‘force of infection’) for the *n*-th individual within the *j*-th age bin, where one initially draws human predispositions to worm uptake from a unit-mean and 1/*k*_*j*_-variance gamma distribution^1^

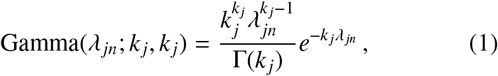

and one evolves the contribution to the force of infection in the *j*-th age bin according to the following differential equation

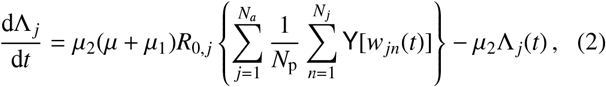

where it is important to note that the quantity which accounts for egg input into the reservoir from sexually-reproducing worms

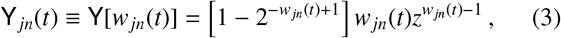

depends on the *total* number of worms *w*_*jn*_ (as opposed to just the number of females) in the *n*-th individual within the *j*-th age bin. Throughout we shall assume that the standard exponential relationship for STH which takes into account the density dependent fecundity of worms *z* ≡ *e*^−γ^ [7, 11, 33] is set to γ = 0.08, unless otherwise stated.

Now let us build a corresponding Poisson process in the opposing direction to model the worm and human deaths, given by rates *µ*_1_ and *µ*. In such a model, the total mean worm burden *m*(*t*) is defined as a weighted sum over the mean worm burdens within each age bin *m*_*j*_(*t*) (with *N*_*a*_ age bins in total) — which itself is the average over the individual worm burdens

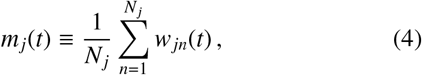

and can hence take different values, depending on the realisation. The stochastic jump equation, which governs its time evolution over a population of *N*_p_ people, takes the following form

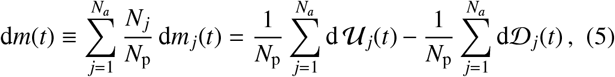

where the worm uptake 𝒰_*j*_(*t*) and (worm and human) death 𝒟 _*j*_(*t*) are given by the following Poisson processes summed from *n* = 1 to *N*_*j*_ individuals within the *j*-th age bin

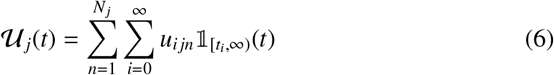

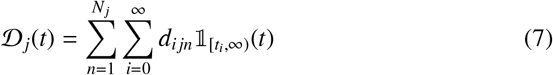

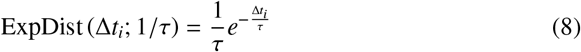

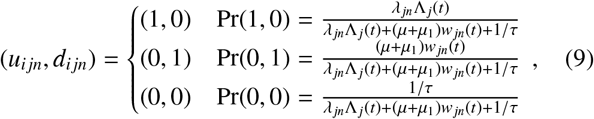

where Δ*t*_*i*_ ≡ *t*_*i*_ −*t*_*i*−1_, τ is a pre-specified timescale short enough such that no event is expected to take place.

At this point, it is important to remark that the stochastic acquisition of new worms per unit time is assumed to follow a 1-step Markov transistion in the model defined above. The basic biological rate of worm establishment within hosts is still unknown, and hence, it is possible that some (if not all) of the clumping pattern observed in worms within hosts may arise from a compounding of the uncertainty in the precise time of worm acquisition with the uncertainty in the number of worms actually acquired by the host in these acquisition events themselves [12].^2^ Hereafter, we leave this as a modelling assumption to investigate further in future work.

### 2.1. Our computational implementation in brief

We have developed a new stochastic simulation code for helminth transmission based on the model defined above. This code comes in the form of a publicly-available open-source python class: [helmpy](https://sites.google.com/view/helmpy), which includes features to incorporate any population stratifications, models of control with MDA, inter-cluster human migration (building from earlier work in Refs. [34, 35]) and an interactive notebook tutorial for calculating all of the quantities discussed in this paper.^3^ This code also includes methods for the stochastic individual-based simulation of schistosome transmission, and is intended to eventually include other helminth infections, such as LF.

The individual-based stochastic model implemented in the [helmpy](https://sites.google.com/view/helmpy) code follows a similar pattern to the Gillespie algorithm [36], however we also adopt a separate methodology for the mean field expansion (which we shall introduce later), which relies on a multi-dimensional Langevin solver, i.e., numerically evolves many coupled drift-diffusion processes simultaneously — see, e.g., Ref. [37].

The code is written in a (mostly) vectorised implementation of the python programming language. To provide a benchmark for the performance, we note here that a single individual-based stochastic realisation of the code which implements multiple MDA treatment rounds and migration between 40 clusters of 500 people (each cluster corresponding to a different infectious reservoir) requires ∼20 minutes to run for 100 years on a laptop with a 1.7 GHz Intel Core i5 processor and 4GB of memory.

### 2.2. The approximate worm burden distribution

Stochastic individual-based simulations of STH transmission seek to incorporate a range of uncertainties into the population-level dynamics of the host-parasite interaction. It is interesting to note that these may typically be categorised into either one, or all, of three important biological/epidemiological components:

1. Uncertainty in predispositions of people (i.e. the specific chance of a particular group of people having their predispositions to STH infection drawn from a probability distribution), i.e., *finite population uncertainty* induced by specific samples of realisations.
2. Uncertainty in precisely when new infections, resulting in new worm acquisition, or worm/host deaths, resulting in worm loss, occur, i.e., *dynamical uncertainty* about precisely when events occur.
3. Uncertainty in the demographic parameters and initial conditions of the given human community (or ‘system’) under consideration, i.e., either *prior* or *posterior uncertainty* on the initial conditions.

One may average over the above forms of uncertainty either individually, or in combination. In particular, to obtain the dynamics corresponding to deterministic models of STH transmission, one must average over the first two — which we shall hereafter refer to as an ‘ensemble average’. The ensemble average of Eq. (2) may be written as

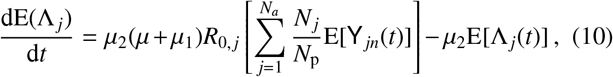

where the result

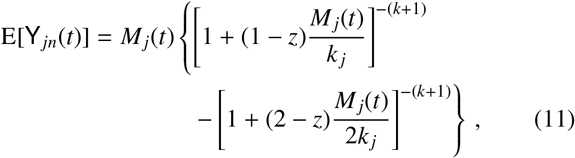

has been derived in numerous Refs. [33, 11, 7]. Note that we have also defined the ensemble average over *m*_*j*_(*t*) as *M*_*j*_(*t*) ≡ E[*m*_*j*_(*t*)]. Performing an equivalent ensemble average over *N*_*j*_*m*_*j*_, the mean-field evolution equation for *M*_*j*_(*t*) is given by

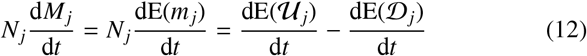

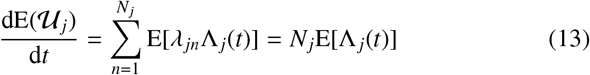

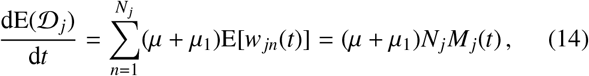

matching the standard theory of the mean or ‘deterministic’ disease model [7]. Note that we have also used Eq. (1) to identify E[λ _*jn*_Λ _*j*_(*t*)] = E(λ _*jn*_)E[Λ _*j*_(*t*)] = E[Λ _*j*_(*t*)] assuming that λ _*jn*_ and Λ(*t*) may be treated as independent random variables. Note that this latter assumption is especially poor when the number of infected individuals (predominately controlled by *k* and/or treatment) is low since a limited number of individuals asserting their influence on the reservoir will lead to stronger cross-correlations of the form E[λ _*jn*_Λ _*j*_(*t*)] −E(λ _*jn*_)E[Λ _*j*_(*t*)] ≠ 0.

In contrast to the simplicity of computing the dynamical equations for *M*_*j*_(*t*) above, calculating the time evolution of *V*_*j*_(*t*) ≡Var[*m* _*j*_(*t*)] will require knowledge of the overall distribution.

It is straightforward to derive a master equation which governs the out-of-equilibrium behaviour of the *n*-th individual’s (in the *j*-th age bin) worm burden distribution as a function of time *P*(*w*_*jn*_, *t*). This is

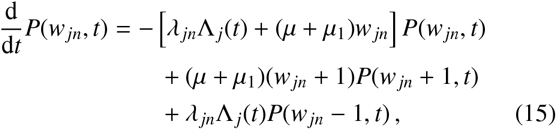

where one recognises the solution as an inhomogeneous Poisson walker Pois[*w*_*jn*_(*t*); ℐ _*jn*_(*t*)] with intensity (for the explicit derivation, see Appendix A)

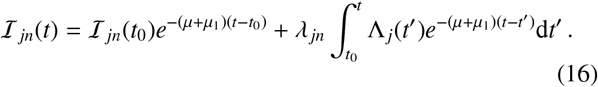

Therefore, for an ensemble of *N*_*j*_ independent walkers (hence individuals) the corresponding probability mass function is also that of a Poisson distribution

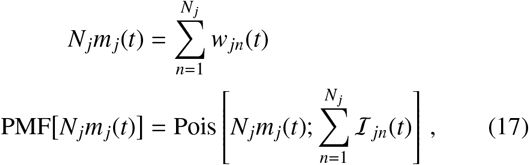

where we have also used the definition of the mean worm burden in the *j*-th age bin given by Eq. (4). Note that in the case where Λ _*j*_(*t*) is roughly constant for all time, the timescale for the distribution to achieve stationarity is Δ*t*_stat_ ≃ 1/(*µ* + *µ*_1_).

Note that by assuming that the reservoir of infection Λ(*t*) in Eq. (16) is deterministic in time and averaging over λ _*jn*_ according to Eq. (1), one recovers the well-established [7] negative binomial distribution for worms within hosts as the result of a gamma-poisson mixture. In reality, of course, the sample mean estimate for the number of infectious stages entering the reservoir in Eq. (2) will induce stochastic fluctuations in Λ(*t*). Estimating the amplitude of these fluctuations will be essential to evaluating the distribution of worms within hosts in the stochastic individual-based simulation.

To begin with, we must first understand the variation in Λ _*j*_(*t*) that is induced from the finite sample of *N*_*j*_ individuals assigned with an initial value of λ _*jn*_. The implicit solution to Eq. (2) is given by

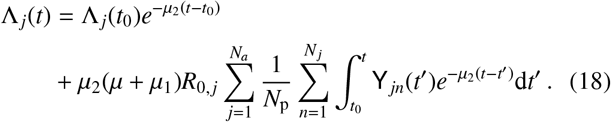

Between ensemble realisations of Eq. (18), the only source of random variation is the sample variance in individual worm burdens when computing the value of the double sum

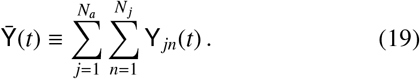

As discussed in Ref. [35], the extremely short reservoir timescale 1/*µ*_2_ compared to the timescales of the other processes, e.g., 1/*µ* and 1/*µ*_1_, allows for an accurate approximation to Eq. (18) to be made where one may coarse-grain (integrate) over time such that

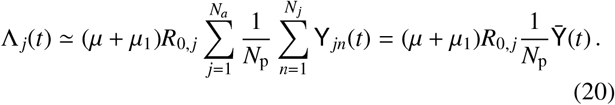

If one now also assumes that *w*_*jn*_ is drawn from an approxima_*N*_tely negative binomial distribution then the distribution of 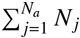 samples drawn from the individual’s egg input distribution is approximately also a negative binomial with updated mean and variance [35] such that

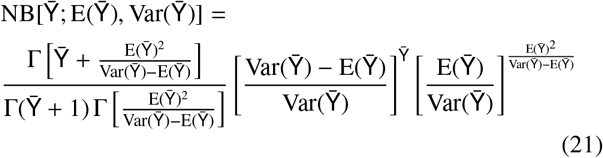

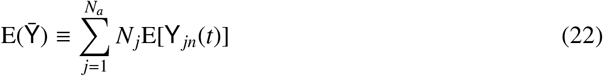

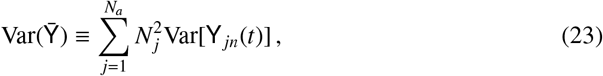

where one may compute the variance of the egg input distribution through evaluation of its second moment, yielding

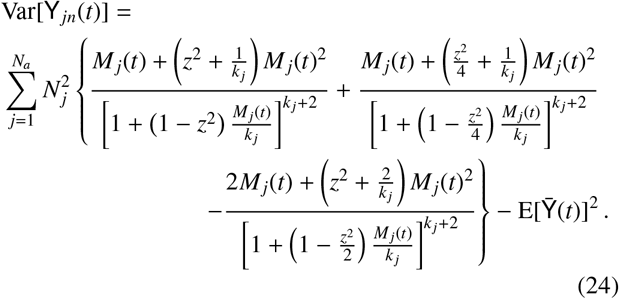

By combining Eqs. (1), (17), (20) and (21) we may infer that the full distribution over ensemble realisations of *P*(*w*_*jn*_, *t*) may be approximated by

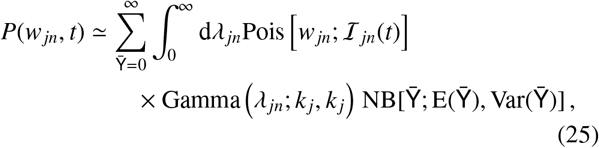

where we have marginalised over all possible reservoir configurations 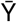 (within the negative binomial approximation) and individual uptake rates λ _*jn*_. In Appendix B we calculate the approximate ensemble mean and variance of the mean worm burden constructed from summed samples from Eq. (25). Sampling directly from Eq. (25), one may effectively reconstruct an accurate ensemble of realisations at any specified point in time as long as the negative binomial approximation of the reservoir is accurate. However, accurate temporal correlations between the sum of individuals will require alternative methods.

### 2.3. A mean field expansion of the system

In order to obtain a better approximation to the temporal correlations of the system, we shall perform a mean field expansion which allows us to calculate an accurate approximation to the stochastic noise around the mean worm burden of the deterministic STH model. Let us now rewrite the value of *N*_*j*_*m*_*j*_ in separate components which depend differently on the system size *N*_*j*_, to obtain

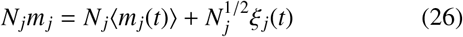

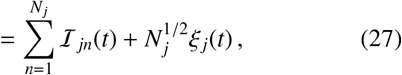

where *ξ* _*j*_(*t*) denotes a fluctuation in each age bin at each moment in time.

Note that in Eq. (26) we have performed a *temporal* average (as opposed to ensemble average) over the states of the Poisson walkers, each with intensity ℐ_*jn*_(*t*) — see Eq. (17). The full ensemble of realisations for the system, which includes a random set of values chosen for λ _*jn*_, is not strictly ergodic. Hence, in order to assess the temporally correlated behaviour, in this section we shall use the temporal average (with notation) to refer to drawing the random values of λ _*jn*_ *a priori* for a given realisation of the system — averaging over only the uncertainty component 2. as discussed at the beginning of Sec. 2.2. The full ensemble may then be restored subsequently by a second averaging over finite population samples (component 1. of Sec. 2.2) *a posteriori*.

In Appendix C we perform a van Kampen [38] expansion to obtain a Fokker-Planck equation for the evolution of the distribution over fluctuations *ξ* _*j*_ in time, which is

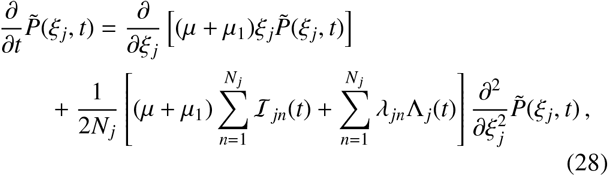

which has the following stationary solution

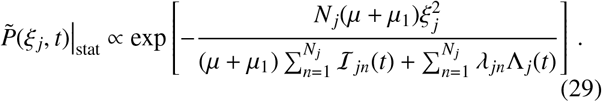

The exact form of the force of infection Λ _*j*_(*t*) is not generally known, however in the limit where the mean-field equations apply, the following estimator may be intuited which uses the ensemble average

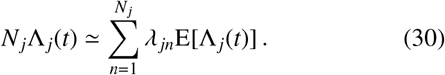

Using the approximation in Eq. (30) we may thus use the solution to the mean field Eqs. (10) and (12) to stochastically evolve trajectories of the mean worm burden with temporal correlations.

### 2.4. Deterministic models versus stochastic simulations

Having established the main mathematical preliminaries, in the analysis of our results it will be convenient to refer to specific configurations of the human heminth system (according to initial choices of transmission parameters) to illustrate our results more effectively. These configurations shall always include the following choices corresponding to the hookworm timescales of: the adult worm death rate *µ*_1_ = 1/2 per year (or an average life span of 2 years); the eggs/larvae death rate in the infectious reservoir *µ*_2_ = 26 per year (or an average life span of 2 weeks) and a density-dependent worm fecundity power of γ = 0.08 [39]. We also have provided the particular choices of parameters in the configurations C1 and C2 of Table 1. Configuration C1 has been chosen with parameters such that the system is far from the unstable ‘breakpoint’ equilibrium as illustrated in Fig. 1, conversely, configuration C2 has been chosen such that the system is much closer to the breakpoint for a clear comparison.

**Table 1:** The particular configurations with hookworm parameters [39] used for demonstration in this work.

| Configuration | Parameter choices ($t_0$ is initial time) |
| --- | --- |
| C1 | $R_0 = 3.5$; $k = 0.3$;<br>$M(t_0) = 2.9$; and $\Lambda(t_0) = 1.25$ |
| C2 | $R_0 = 2.1$; $k = 0.5$;<br>$M(t_0) = 2.1$; and $\Lambda(t_0) = 1.1$ |
| All cases | $\gamma = 0.08$; $\mu = 1/70$; $\mu_1 = 1/2$;<br>and $\mu_2 = 26$ (last three are per year) |

In Sec. 2.2, we stated that deterministic models of STH transmission are obtained by averaging over both of the uncertainties in predispositions to infection and when events precisely occur in time. As such, the first of our results regarding the relationship between the deterministic models of STH transmission and stochastic simulations is not unexpected. The deterministic predictions for the dynamics of the mean worm burden [7] and the *mean of the mean* worm burdens derived from individual-based simulations exactly match — see, for instance, the good match between dashed lines plotted in the left and right top panels of Fig. 2, where we have plotted the dynamics of the mean worm burdens in a community with parameters in configuration C1 (see Table 1) for a range of population sizes. The left and right columns of this figure correspond to the numerical solution to the fully individual-based stochastic simulation that we defined at the beginning of Sec. 2 and the mean field model with the stochastic noise approximation using Eqs. (27) and (28), respectively.

**Figure 2:**
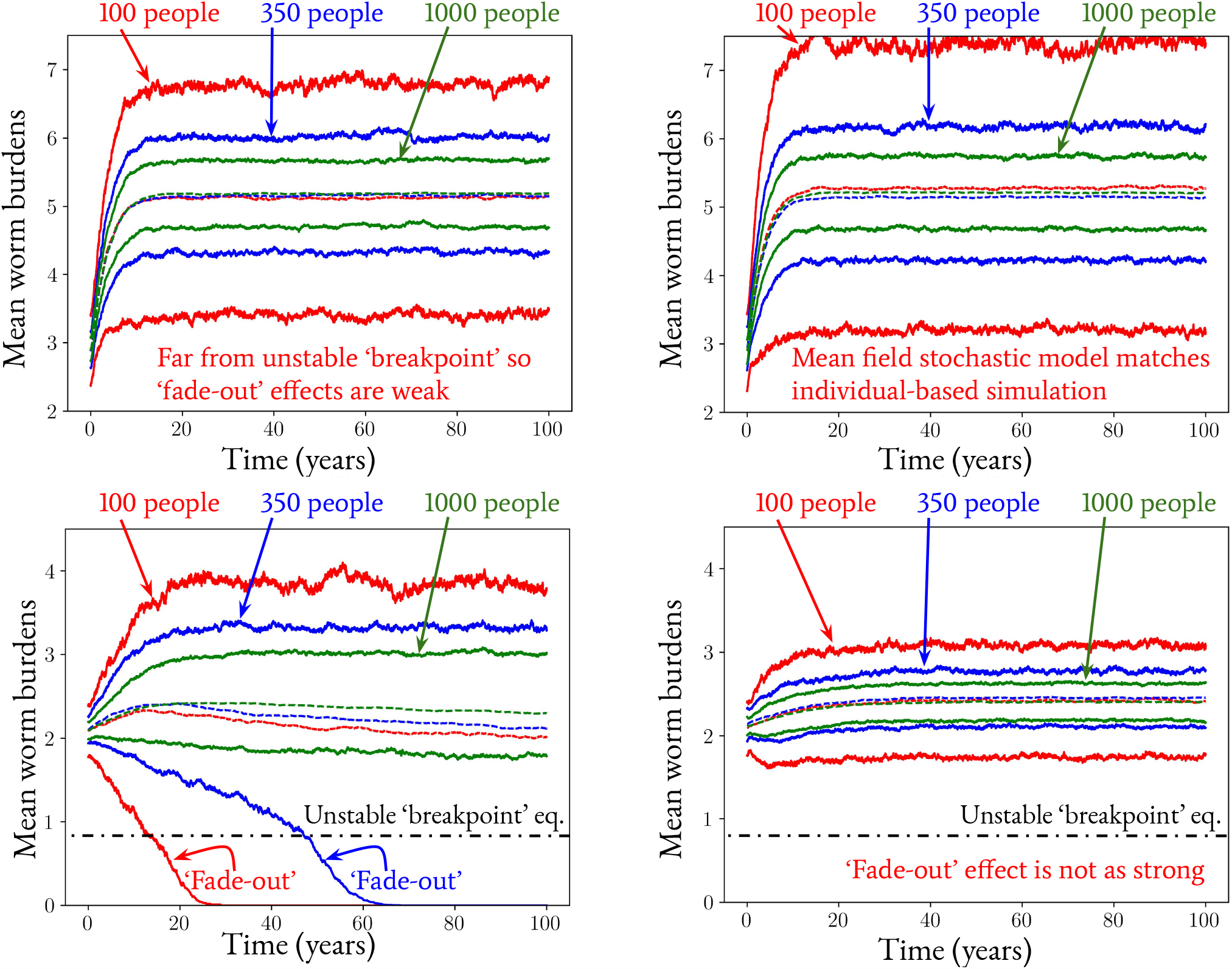
Two different examples (in both left and right panels) of the expected (dashed lines) and 68% credible regions (between the solid lines — used due to the overlap with the standard error for the normal mean) of the possible mean worm burdens realised in time by the stochastic fully individual-based simulation method (left column) and the counterpart simulations using a mean field stochastic approximation method (see Eqs. (27) and (28)) have also been run for comparison (right column). The top row corresponds configuration C1 in Table 1. The bottom row corresponds configuration C2 in Table 1. In each case 250 realisations were run to construct the statistics.

Considering the same pair of plots (top row) in Fig. 2 in more detail we find that, for a given range of parameters, the two methods used to generate the 68% credible intervals about each mean value also agree to very good accuracy. This agreement is not always the case, however, as we illustrate with the same pair of plots but with different transmission parameters on the bottom row of Fig. 2. Given configuration C2 (see Table 1), in which the endemic equilibrium value is situated closer to the unstable ‘breakpoint’ equilibrium, we find that an important phenomenon which exists in the individual-based stochastic simulation is not present in the deterministic model — this phenomenon is known as ‘fade-out’ in the parasite population. ‘Fade-out’ is where transmission is interrupted due to chance events, even when the underlying transmission success (the value of *R*_0_) is above the level that deterministic models predict will result in parasite persistence. In other words, values just above the transmission breakpoint of the deterministic model may indeed move to parasite extinction, due to chance effects.

Ordinarily in disease transmission models, the stochastic noise approximation using Eqs. (27) and (28) may be used to compute similar ‘fade-out’ effects to great accuracy, see, e.g., in an SIR model [40]. However, the presence of the unstable ‘breakpoint’ equilibrium leads to a stronger fade-out effect as one can observe in the drift towards a zero mean worm burden (transmission interruption) of the lower 68% credible interval contour (and mean) in the bottom left plot of Fig. 2 with population sizes of 100 and 350, and which markedly reduces the accuracy of the mean field stochastic approximation. A simple Gaussian approximation notwithstanding, fade-out is a very important phenomenon of considerable practical relevance to the prospect of helminth transmission interruption. Note that the probability of a fade-out event also increases from very small to large as village population size decreases, as is also shown by the results of Fig. 2. The inability of the stochastic noise approximation model to capture the fade-out phenomenon is of considerable interest and requires a more detailed mathematical description of the stochastic dynamics of the reservoir of infection. Such work is planned for investigation in future work.

To illustrate how the strength of the fade-out phenomenon we have described is influenced by the human-helminth system proximity to the unstable equilibrium, we have included Fig. 3. This plot corresponds to the same simulated system as shown in the lower left panel of Fig. 2 (configuration C2), but where the initial ensemble mean worm burden has been halved to *M*(*t*_0_) = 1.05. From this plot it is immediately clear that as the system nears the unstable breakpoint, the strength of fade out increases — as illustrated by the much more rapid descent of the lower lines demarcating the 68% credible regions and the strong fade out even with 1000 individuals (the green lines).

**Figure 3:**
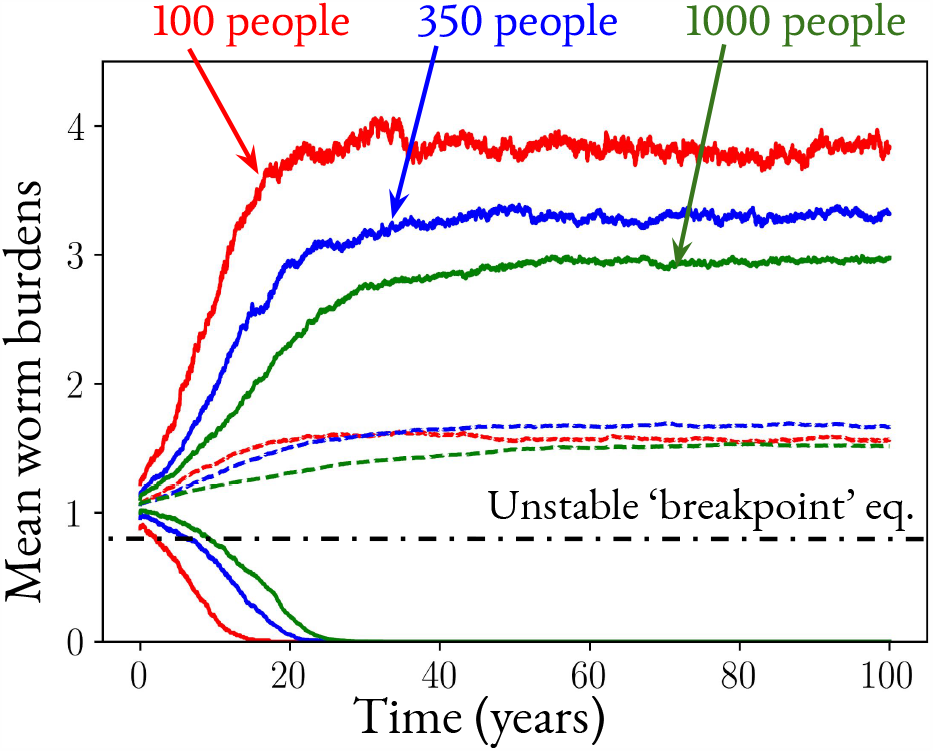
The equivalent stochastic simulations to the lower left panel of Fig. 2 (configuration C2) but with half the initial ensemble mean worm burden *M*(*t*_0_) = 1.05.

In our analysis in this section, although we ignored age structure, its inclusion does not influence the general insights although it will influence quantitative detail. For hookworm, the absence of age structure is not too serious, even in quantitative terms since field studies suggest that the force of infection is often independent of age [39]. To illustrate this point, in Fig. 4 we have plotted the C1 configuration (see Table 1) with relevant hookworm age stucture included, where the difference between Fig. 4 and the top left plot of Fig. 2 is very small.

**Figure 4:**
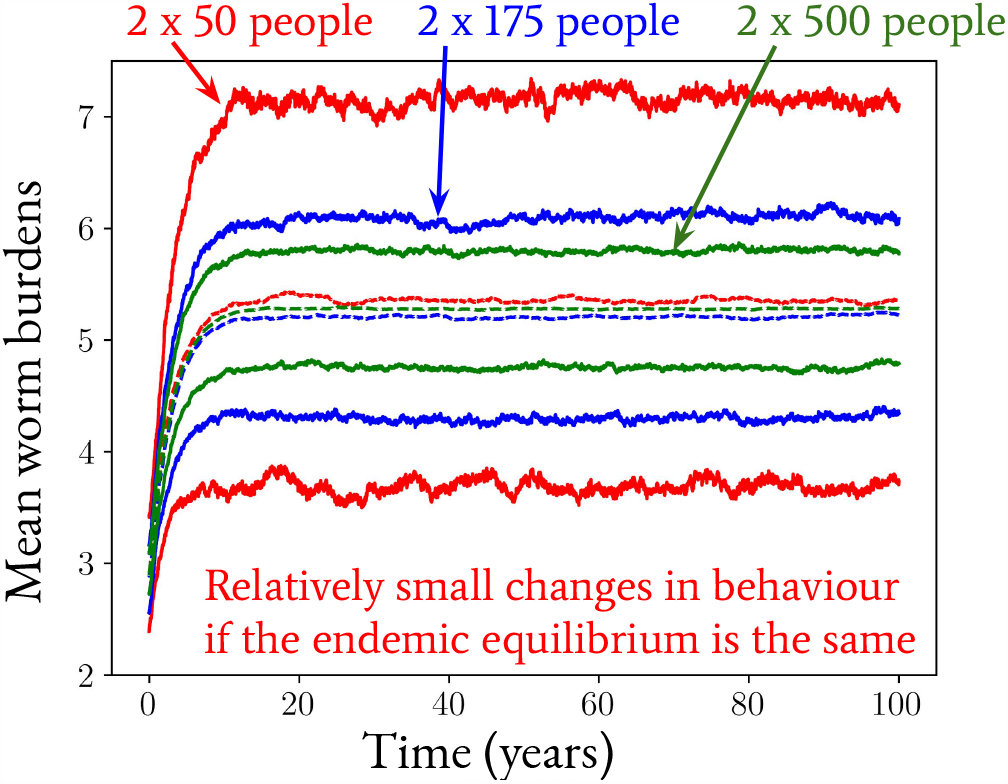
An example demonstrating the relatively small effects of age structure on stochastic individual-based model with the original configuration of C1 in Table 1. In this instance, the population has been split into two equal-sized components of ages 0-14 and 15+, where we the worm uptake rate in the case of the 15+ group has been set to exactly 3 times that of the 0-14 group. The observed lack of an effect may be restricted to hookworm infections where the force of infection is often constant across age classes. This is not the case for *Ascaris* and *Trichuris* infections [26]. In each case 250 realisations were run to construct the statistics.

### 2.5. The ‘breakpoint’ of stochastic simulations

STHs are dioecious and therefore both male and female parasites must be in the same host for the female worm to be fertilized and produce viable eggs or larvae which sustain transmission. Past research in the context of the deterministic models of transmission has demonstrated that there are three apparent equilibria: a stable endemic state, parasite extinction, and an unstable state termed the ‘transmission breakpoint’ which lies between the stable state and parasite extinction [41, 42, 43]. The existence of such a breakpoint can be intuited by the above limitations of STH reproduction when it becomes difficult to find a male and female worm pair within an individual host and it represents a clear target for control policies which aim to achieve transmission interruption. As we also discussed earlier in Sec. 1 and illustrated in Fig. 1, this (dashed) breakpoint curve in the phase plane of Fig. 1 acts as a separatrix between the attractor basins of the stable (endemic) and extinction equilibria.

Extensive past numerical analyses based on using the individual-based simulation code [44, 32] strongly indicate that the uncertainties which are inherent in individual-based simulations dominate the transmission dynamics near the deterministic breakpoint in transmission. This is especially true if the breakpoint is close to the endemic equilibrium of parasite extinction (as opposed to the stable equilibrium of endemic infection) as a consequence of high degree of parasite aggregation in the human population [45, 7], as is illustrated by the appearance of the fade-out effect between the top left and bottom left plots of Fig. 2. Furthermore, in the previous section, we highlighted some limitations in the standard formulation of deterministic models to account for the specific phenomena of population ‘fade-out’ (or spontaneous transmission interruption) which are present in the real world due to chance effects in population growth and decay especially when host population densities are low. Such effects are also captured by individual-based stochastic simulation models. It may hence be reasonable to ask whether concepts which exist in the deterministic framework are present in the stochastic simulations.

Conclusions drawn from hookworm simulation studies in Refs. [32, 44] for a range of population sizes study thresholds in prevalence in the range 0.5%-2% below which the probability of transmission interruption is assessed. In particular, attaining a prevalence below the threshold of ∼1% for population sizes 100-1000 leads to interruption of transmission with a high probability. In this section, we provide a theoretical argument for the existence of this threshold as a ‘stochastic breakpoint’ which works in a similar way to the behaviour present in deterministic models, but accounting for some unavoidable uncertainty.

Let us define a net ‘grower’ as an individual who, given a particular reservoir of infection and adult worm death rate, is able to accumulate more than one worm over time in a consistent manner (an individual who is predisposed to infection). Such individuals are crucial to the survival of the parasite in a defined community as they provide future material to the reservoir of infective stages in the habitat which subsequentially may grow the number of human hosts who become net growers of infection themselves, and so on. One can typically state that if the number of human hosts in which the parasite population grows (net growers) is one or more in a given cluster or community of people, then the chances of parasite extinction in the long-term are low.

Immediately after, e.g., treatment through an MDA programme, only a limited number of infected individuals remain in each age bin *N*_inf_ and hence are able to contribute to the reservoir in the following timesteps (until further relaxation of the system towards some equilibrium). Using a modified version of Eq. (20) we may infer the following estimate of the infectious reservoir contribution by those individuals into the *j*-th age bin

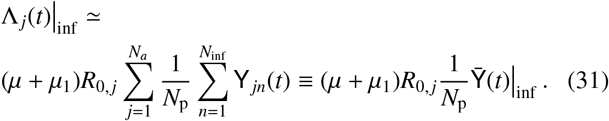

Hence, by further assuming that the fluctuations in the reservoir of infection are well-approximated by the negative binomial distribution, we may evaluate the expected maximum fraction 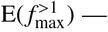 — of all individuals in the cluster — whose ratio between their uptake rate λ _*jn*_Λ _*j*_(*t*)|_inf_ and the death rate of a single worm (*µ* + *µ*_1_) exceeds 1 through the following integral motivated by the form of Eq. (25)

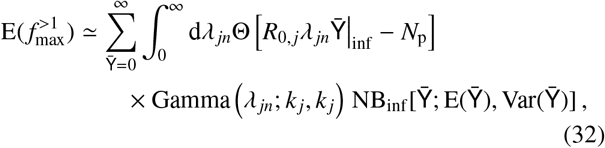

where 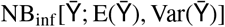 denotes the reservoir egg count negative binomial distribution corresponding to 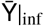 and Θ(·) is a Heaviside function. Given Eq. (32) the expected number of net growers is therefore simply 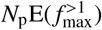.

Let us now define the prevalence of infection p as the fraction of the total population who are infected. We therefore deduce that p = *N*_inf_ /*N*_p_. Using Eqs. (25) and (32), we have plotted a calculation of both the probability of the number of net growers for communities with configurations C1 and C2 (see Table 1) in Fig. 5 and the expected number of net growers as a function of the prevalence in Fig. 6 for a wide range of system configurations. In both sets of plots we consider population numbers of 100, 350 and 1000, as indicated.

**Figure 5:**
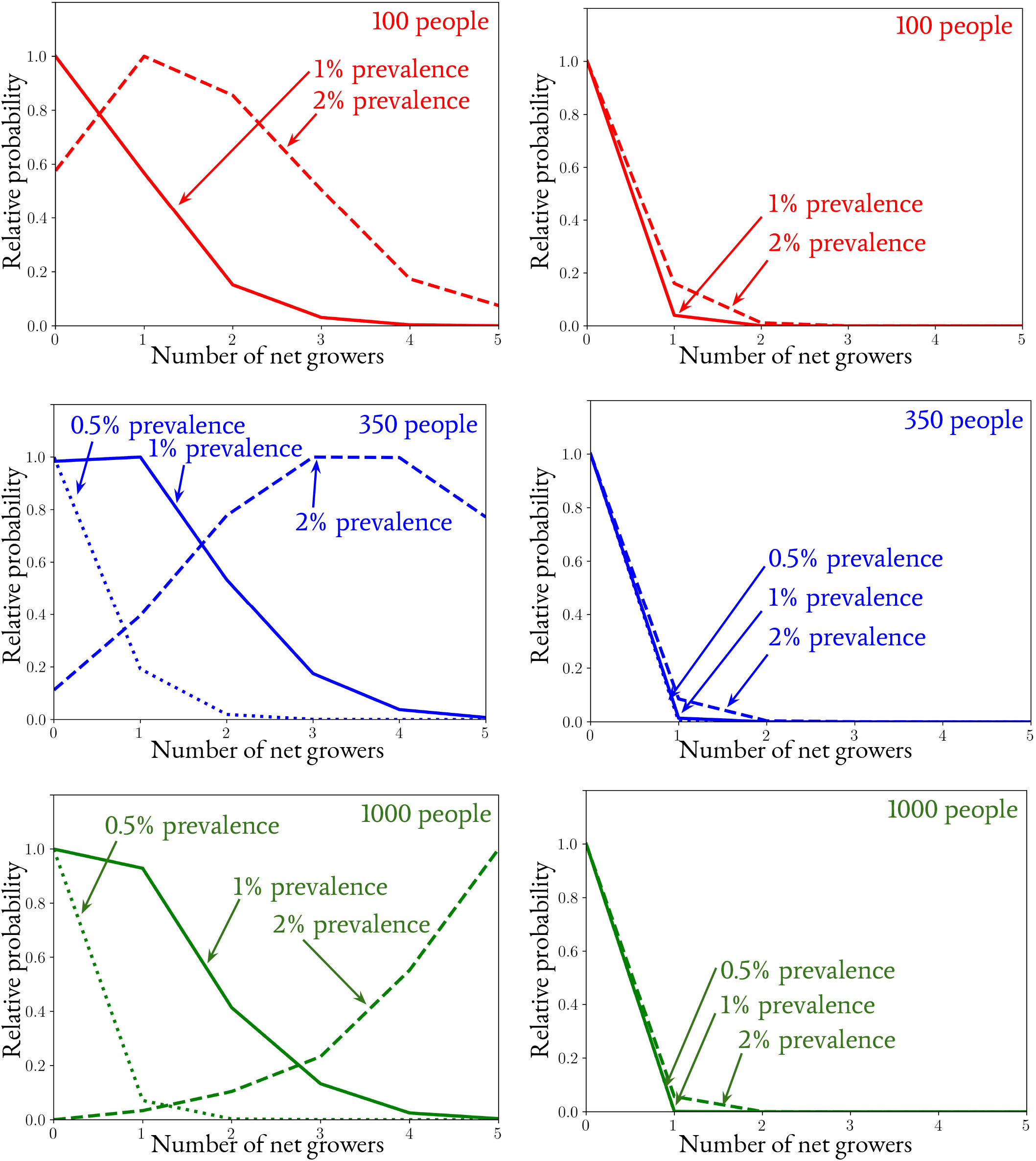
The relative probability (the probability mass divided by the maximum value) of a number of individuals who will gain more than one worm, or, ‘net growers’ given that effectively only one individual is contributing to the reservoir of infection per unit time for two different system configurations (given by the left and right columns). This probability has been calculated using Eq. (25) and the condition for a net grower — as discussed in Sec. 2.5. The left columns have the same parameters of the system as those of configuration C1 in Table 1 (which is assumed to have attained an endemic equilibrium mean worm burden). The right columns have the same parameters of the system as those of configuration C2 in Table 1 (which is also assumed to have attained an endemic equilibrium mean worm burden).

**Figure 6:**
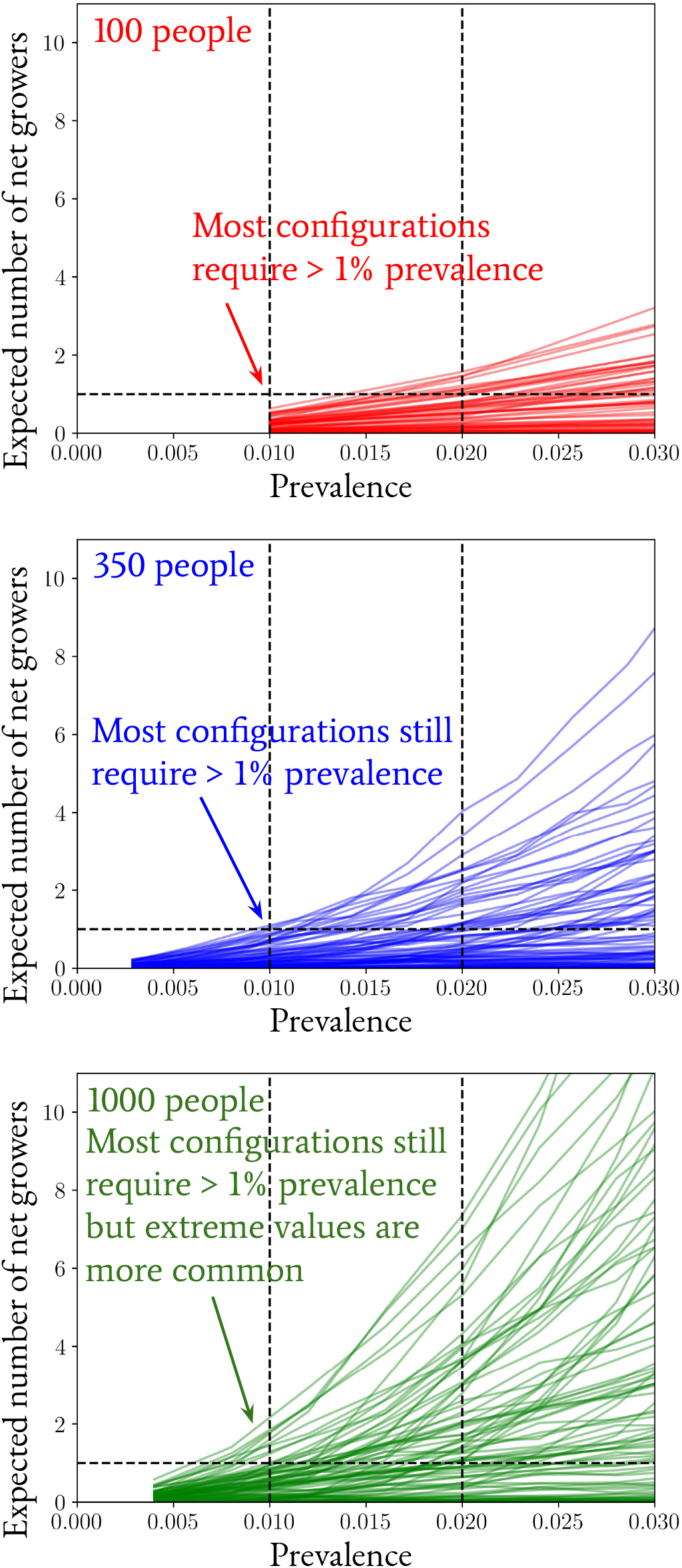
The expected number of individuals who will gain more than one worm, or, ‘net growers’ given that effectively only one individual is contributing to the reservoir of infection per unit time for a range of different system configurations. These values have been obtained by randomly sampling configurations of Eq. (32) with flat priors defined over the following intervals in one cluster without age structure for illustration: the basic reproduction number *R*_0_ ∈ [2, 4]; the logarithim (base 10) of the worm aggregation parameter log_10_ *k* ∈ [−4, 0]; the initial mean worm burden *M*(*t*_0_) ∈ [2, 5] and an assumed density-dependent worm fecundity power of γ = 0.08. The range of system configurations sampled corresponds to much wider range than those given in Fig. 5.

Fig. 5 demonstrates that for system configurations further from the unstable ‘breakpoint’ equilibrium — configuration C1 in the right column of plots — the probability of exceeding a net number of growers larger than 1 becomes large once the system exceeds the 1% prevalence threshold, with the contrary being true below this threshold. The plots in the left column — which correspond to configuration C2 — the probability for the number of net growers to be above 1 remains low even for a prevalence of 2%, which is to be expected due to this configuration’s proximity to the unstable ‘breakpoint’ equilibrium. Furthermore, from Fig. 6, it is immediately apparent that the expected number of growers is extremely unlikely to exceed 1 if the prevalence of infection is lower than ∼1% in all cases of population number. Hence, this threshold essentially acts as a ‘stochastic breakpoint’, below which the probability of interrupting transmission in stochastic individual-based simulations becomes very likely.

Note that caveats to our analysis here include varying the population number well beyond the prior limits used in our short sensitivity analysis — in such situations it is possible for apparent thresholds to go much lower or higher, e.g., consider the small, but non-negligible probabilities of sustaining transmission below a prevalence of 1% for much larger population numbers than those studied here in Ref. [44]. It is important to also mention that the approximate bound we obtain on the prevalence as a ‘stochastic breakpoint’ applies as more of a strict bound and hence does not exclude the possibility (or even high likelihood in some specific configurations) of elimination at a higher prevalence of 2%. Our discussion here is therefore complementary (and not contradictory) to, e.g., Ref. [32].

We expect that similar conclusions apply in settings where parasite interruption is predicted since the basic reproduction number (*R*_0_) is too low to sustain transmission (see Fig. 1). If the mean worm load is just below this critical *R*_0_ value to sustain transmission in regions where the only stable state is parasite extinction, there will be a probability distribution of situations where parasite persistence occurs for a while due to chance events which may obscure the precise location of this threshold.

Our short analysis in this section has provided some theoretical justification for the existence of a ‘stochastic breakpoint’ which has been identified to be an important target for transmission interruption from previous hookworm simulation studies [32, 44] and our analysis may also be extended to other parasites, which shall be subject of future work.

### 2.6. A note on modifying the reservoir to account for ageing

In the stochastic model we have built, the effect of people ageing over time has been neglected so far. Assuming that the effective number of people in each age bin remains unchanged, the passing of individuals between age bins may be fully accounted for as a reservoir pulse of the same form as Eq. (33) which introduces new egg pulses drawn from the (*j* −1)-th age bin and removes eggs from the *j*-th bin into the (*j* + 1)-th bin.

The reservoir fluctuation model described above demonstrates how ageing can can only affect the system by a significant variation in *k* _*j*_ between bins — furthermore, the influence of such fluctuations on the reservoir of infection can be shown to be damped significantly when the age bins are much wider than 1/*µ*_1_ due to the relaxation timescale of each individual Poisson walker (see, e.g., Eq. (17)). In light of this fact, we have elected to avoid a direct description of ageing in this work without loss of generality in our argumentation, while leaving future versions of our stochastic simulation to include these sub-dominant effects. Past published individual-based stochastic models of helminth transmission have included a dynamically ageing population [26].

## 3. Extensions to the stochastic individual-based model

### 3.1. The effect of infected human migration

Up until this point we have not considered processes which may modify the standard STH transmission model. Applying control measures through MDA and the effect of infected human migration between clusters are both processes which modify the dynamics. The impact of MDA has been covered in various recent publications [46, 32, 44], whereas, infected human migration in the context of STH transmission has only recently been addressed in the context of its impact on achieving breakpoints in transmission [34, 35]. In the context of STH infections it is necessary for infected people to be present at a location long enough to contribute to the infectious reservoir.

In this paper, we incorporate the migration model for STH described in Ref. [35] into our individual-based stochastic simulation. We shall use this model to describe the consequences that infected human migration has on human helminth transmission using illustrative examples. In Ref. [35] the relative size of the migration rate of individual egg counts into or out of a region (e.g., some number of people per year) compared to the death rate of infectious stages within the reservoir, *µ*_2_ (*µ*_2_ = 26 per year for hookworm), was found to be have important implications for the effect of migration on transmission. Hence, in all of the results on migration in this section, the migration rate shall be quoted as a factor of *µ*_2_ to investigate the importance of the ratio between these two parameters in the fully individual-based stochastic simulation.

The migration of infected individuals between clustered communities represents a combination of both the *finite population* and *dynamical* uncertainty described at the beginning of Sec. 2.2. A simple modification to the model we have introduced, which builds from the work in Ref. [35], may be considered to incorporate the effects of human migration into the transmission dynamics: consider a compound Poisson process t _*j*_(*t*) which perturbs Eq. (2) in the following way^4^

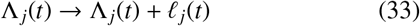

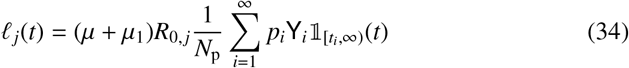

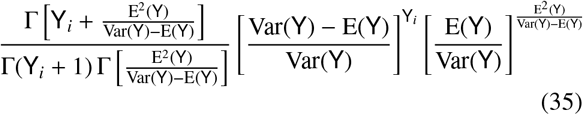

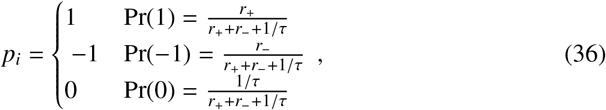

and where *r*_+_ and *r*_−_ are the ingoing and outgoing the migratory rates, respectively, and we have coarse-grained in time to obtain the t _*j*_(*t*) fluctuation amplitude with respect to the mean worm burden — as in Eq. (20). With the effect of migration defined in this way, the parameters of Eq. (35) may be randomly selected from any of the corresponding available age bins, and the ensemble mean updated to account for any out-of-equilibrium behaviour. Using a similar set of pulses, in Ref. [35] it was further shown that the migratory rate must exceed or be around the same order as the death rate of infectious material in the reservoir, i.e., *r*_+_, *r*_−_ ≳ *µ*_2_, for the migration to affect the dynamics significantly.

In Ref. [35] it is also assumed that the reservoir pulses which arise from migration are uniform across the full ensemble of realisations: though this captures the inherent variability due to migration effects, it does not marginalise entirely over the full ensemble of possible fluctuations which are captured in this work. It is necessary in the finite population limit, then, for such processes to be modelled by a full stochastic individual-based simulation, where we leave the possibility of an analytic description for future work.

In light of the apparent necessity for a numerical approach, we also point out that when using a fully individual-based simulation one may randomly draw from the worm burdens of the individuals and compute an egg pulse amplitude using Eq. (3) —in doing so potentially capturing the out-of-equilibrium behaviour of the reservoir during, e.g., rounds of treatment. For the latter reason we shall adopt this approach when generating our numerical results throughout. We further note that such an approach has also been advocated in similar models such as those in Ref. [34].

In the bottom left panel of Fig. 2 we provided an example of the fade-out effect which appears in standard individual-based simulations. In metapopulation disease transmission models it is well-known that migration effects can stabilise local tranmission, evading effects such as fade-out even when the population sizes are small — see, e.g., Ref. [47]. By including migration from a cluster with configuration C1 (see Table 1) into the cluster depicted in the plot with configuration C2 (with 350 people each), in Fig. 7 we demonstrate that a similar stabilisation of the fade-out effect can also occur for helminth transmission models, despite the especially strong fade-out induced from the presence of the unstable ‘breakpoint’ equilibrium. As targets for helminth elimination trials drive the prevalence in regions closer to (and beyond) the unstable equilibrium, this is an important possibility to take note of with practical implications should migration within a given region be of the same order or higher as is indicated in Fig. 2. Note that in order for this effect to be important, the migration rate needs to exceed the death rate of eggs/larvae in the infectious reservoir, i.e., *r*_+_, *r*_−_ ≳ *µ*_2_, which is an early confirmation of the conclusions drawn from Ref. [35], but we shall now investigate the importance of migration rates relative to *µ*_2_ in more detail by examining another new consequence of migration.

**Figure 7:**
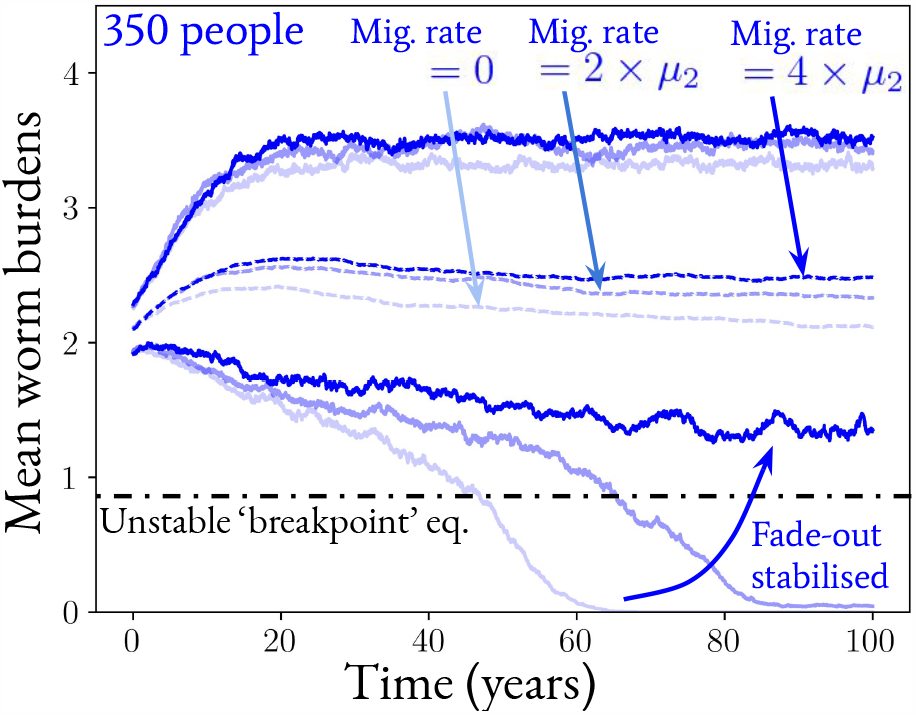
Three different examples (in both left and right panels) of the expected (dashed lines) and 68% credible regions (between the solid lines — used due to the overlap with the standard error for the normal mean) of the possible mean worm burdens realised in time by the stochastic individual-based simulation method with the inclusion of infected human migration from a cluster (of 350 people) with the same transmission parameters as configuration C1 in Table 1 into a cluster (of 350 people) with the same transmission parameters as configuration C2 in Table 1, the latter of which is displayed. Three different migration rates of individuals are used as scales of the death rate of eggs/larvae in the infectious reservoir *µ*_2_ and are depicted by different shades of blue colour. In each case 250 realisations were run to construct the statistics.

Note that in Fig. 7 we have chosen the population size of 350 individuals to illustrate the stabilisation of the fade-out phenomenon since this provides a nice comparison with the bottom left panel of Fig. 2. As can be inferred from the latter, by increasing the population size used in the same example, the fade-out phenomenon would eventually self-stabilise over the time interval of interest (as it does for our example at 1000 individuals). Concommitant with this fact is that a lower net migration rate into the cluster would be required to aid this stabilisation, however, for significant effects at all to occur we find that migration rates with order of magnitude comparable to, or above, the scale *µ*_2_ are still necessary to induce this effect.

Migration into a cluster may also occur from an untreated external source (UES), i.e., from a location with a defined negative binomial distribution of worms within hosts and population number that is left untreated in the endemic state. In such instances, it is also possible that the region may have previously achieved elimination which is then reversed — an ‘outbreak’ scenario.

In Fig. 8 we have plotted the prevalence of worms within hosts after a period of 100 years for a previously-eliminated cluster experiencing UES migration for a range of migration rates (as a ratio of the eggs or larvae death rate for comparison). The transmission parameters chosen for the UES match a cluster of the indicated population size and configuration C1 from Table 1 from which migrants move to a cluster with the same indicated population size and transmission parameters from configuration C2 in the left column of plots (configuration C1 in the right column of plots), where the latter cluster has been initialised without any initial infections. From these plots one can immediately see that a discrete transition occurs in prevalence after a critical UES migration rate has been reached and that this value changes according to population size. This transition marks the point at which the STH transmission is sustained endemically, and hence the UES migration rate that this corresponds to is of critical importance to policy makers and implementers of STH control programmes since achieving elimination above this critical point in migration rate is likely impossible without treating the external source of migrants simultaneously.

**Figure 8:**
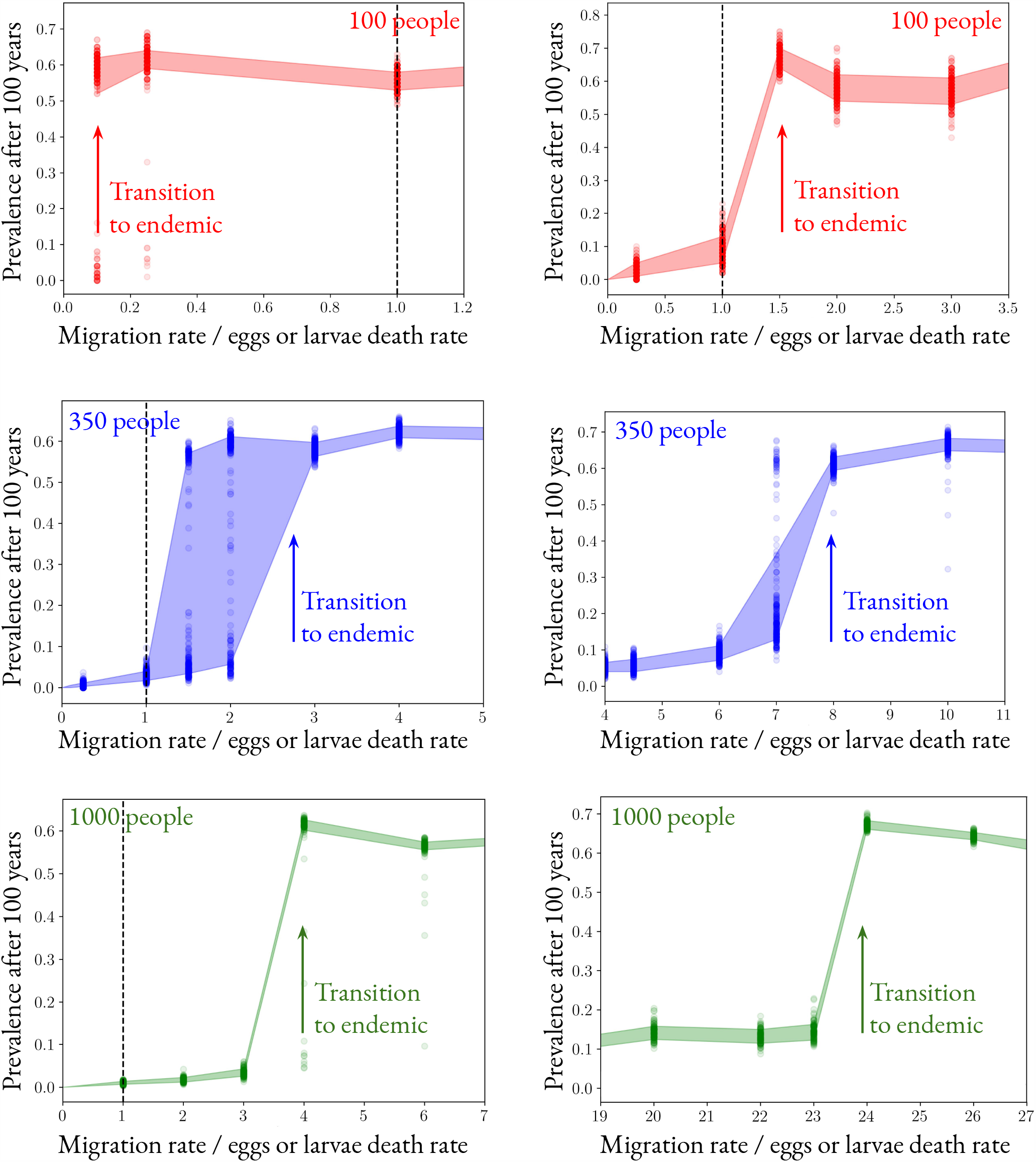
The prevalence of worms within hosts after a period of 100 years for a previously-eliminated cluster experiencing migration from an untreated external source (UES) for a range of migration rates. The transmission parameters chosen for the UES match a cluster of the indicated population size and configuration C1 from Table 1 from which migrants move to a cluster with the same indicated population size and transmission parameters from configuration C2 in the right column of plots (configuration C1 in the left column of plots), where the latter cluster has been initialised without any initial infections. Note that we are using the death rate of eggs/larvae in the infectious reservoir, *µ*_2_, as a unit with which to measure the relative migration rate, where *µ*_2_ = 26 per year for hookworm. Dashed vertical black lines indicate when the migration rate equals *µ*_2_.

For the larger population numbers (350 and 1000), Fig. 8 also confirms the conclusions drawn in Ref. [35] — that critical migration rates to affecting transmission must exceed an order of magnitude threshold of at least *µ*_2_. Note, however that the precise number of individuals required to be contributing to the reservoir per unit time can be substantially modified when the population number is changed and the apparent prevalence required to trigger the outbreak transition can be much higher than 1% — typically 10%. Therefore, it is important that we state that the stochastic breakpoint that we derived in Sec. 2.5 only applies to situations where the prevalence of infection is lowered due to treatment, as opposed to being raised from a zero worm state due to UES migration. Such a difference in breakpoint prevalence value required between these two scenarios is due to the greater homogeneity in worm loads of individuals in the case of UES migration, as opposed to the case of post-treatment where a limited number of individuals have a large number or worms — the latter of which is more consistent with the negative binomial reservoir approximation used to derive the stochastic breakpoint in Sec. 2.5. We plan to investigate this apparent asymmetry in how the stochastic breakpoint applies for individual-based simulations in future work.

Note here that for configuration C1 and a small population number of 100, in Fig. 8 (top right panel) we see that transmission may transition to the endemic state spontaneously with a much lower migration rate than *µ*_2_ — such an effect is due to the additional variance induced by finite population effects and this represents a potentially important caveat to the migration rate threshold we have proposed above.

### 3.2. Accounting for treatment with random compliance

To demonstrate the difficulty in achieving STH transmission interruption with an MDA control programme when migration of infected humans between clustered communities is in place, even when treating all communities at once, in Fig. 9 we plot the positive predictive value (PPV) of elimination after 100 years having reached at least a given prevalence threshold or below (as indicated on the horizontal axis) in a cluster with a C2 configuration (see Table 1) after 3 rounds of treatment (assuming 100% efficacy). Treatment rounds have been applied in years 15, 16 and 17 simultaneously to clusters with configurations C1 and C2 (with population sizes of 350 each), where the dashed black horizontal line at a PPV of 1 shows that elimination is essentially certain in the C2 configuration after treatment if no migration occurs between the two clusters. Once again, however, we find that this PPV drops sharply as the migration rate between clusters is increased in rate relative to the eggs/larvae death rate in the reservoir, *µ*_2_.

**Figure 9:**
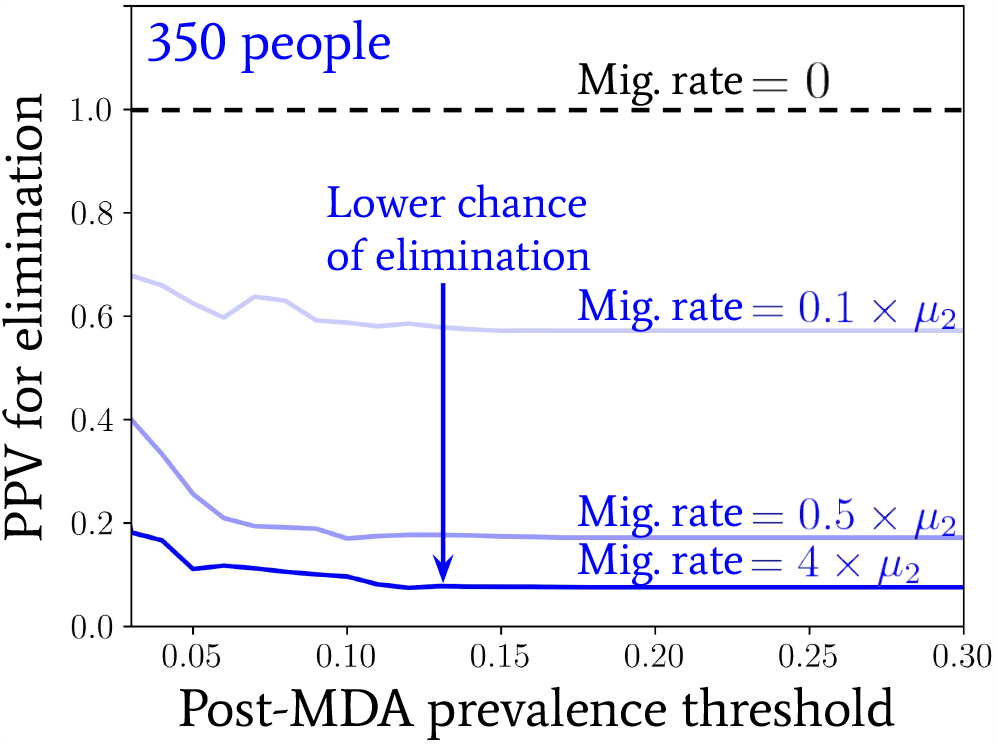
The positive predictive value (PPV) for elimination (the latter is here defined as no remaining worms after 100 years) in a cluster with a C2 configuration (see Table 1) given that a prevalence threshold — which is varied on the horizontal axis — has been crossed immediately post-MDA. In three of the shown cases the C2 cluster is also experiencing migration from another cluster with a C1 configuration (once again, see Table 1) where the migration rates are indicated (using units of *µ*_2_ = 26 per year for hookworm). The treatment has been applied to both clusters and is 3 rounds of MDA with a random compliance pattern of individual behaviours assumed at 60% coverage (and 100% efficacy) in years 15, 16 and 17 of the overall 100.

## 4. Concluding discussion

The control of helminth infections worldwide will require a greater degree of monitoring and evaluation throughout 2020 to 2030 as treatment programmes strive to get to the point where transmission interruption thresholds have been achieved. This leads to an increased focus of research into predicting the likelihood of ‘bounce back’ once the transmission breakpoint has been achieved. Such breakpoints are variously defined, but in general for human helminths, the indication from modelling studies is that a true prevalence (based on the use of a very sensitive diagnostic) of less than 1% moves the dynamics of the system to the interruption of transmission. Defining the break-point and designing monitoring and evaluation programmes requires the use of stochastic individual-based models to determine this likelihood in terms of PPV and NPV values of the probability of transmission elimination or bounce back. It is therefore highly desirable to account for as many sources — natural, or otherwise — of uncertainty to ensure a high degree of confidence in the predictions made by such models. The effect of infected human migration in and out of health implementation units on control outcomes, is but one important example of a key source of heterogeneity.

In this paper, we have sought to answer a number of questions of importance to providing reliable predictions for the design of public health programmes for the control of human helminth infections and their monitoring and evaluation.

Averaged quantities, e.g., the mean worm burden, are matched well by the deterministic model to the individual-based simulations. This is an expected result conformed by past work [12]. Stochastic versions of the deterministic models employing mean-field approximations may be derived to predict how fluctuations in the averaged quantities vary over time. The mean-field approximation, however, appears to fail to predict specific important phenomena accurately which exist in an individual-based simulation; namely, fade-out effects.

Our analyses suggest that deterministic models of STH (and other helminth) transmission provide important general guidelines to making predictions of what level of control, such as MDA coverage, and what patterns of individual compliance to treatment will move the system close to or below the deterministic breakpoint in transmission. However, once in this region, chance effects can result in either transmission elimination (fade out) or bounce back even when above or below the breakpoint. In practice, around the unstable equilibrium, stochastic noise induces a degree of uncertainty in outcome which must be described as a probability of a certain outcome being achieved. Deriving these probability descriptions in terms of, for example PPV and NPV values, requires the use of an individual-based stochastic model.

Our analyses suggest that public health workers will benefit from determining the transmission threshold — or ‘breakpoint’ — for transmission interruption using individual-based simulations, and that theoretical calculations suggest a true prevalence target of below ∼1% to have an extremely high probability of interruption in a wide variety of scenarios for all the major helminth infections of humans. The important public health message, however, is that in a stochastic world where chance events play a central role, nothing is certain. As such, outcomes should be expressed as probability events. Even below the 1% threshold, however, bounce back can occur due to chance events, where its likelihood will depend on host and parasite population size.

Infected human migration, like treatment, is an effect which requires individual-based simulation to assess its importance in sustaining high levels of control in the real world where much heterogeneity exists within and between health intervention population units in most regions of endemic infection. Analyses suggest that sufficiently rapid migration from regions with prevalent infection can shift units in which high MDA coverage is achieved to a situation of sustained transmission. This suggests that external processes such as inter-cluster migration could play an important role in sustaining STH transmission.

The models suggest the existence of an important critical scale (i.e., order of magnitude) for the migration rate (up to the population number caveat discussed in Sec. 3.1) corresponding to the death rate of eggs or larvae in the case of STH infections in the infectious reservoir exists, above which, simulations exhibit various behaviours. These include stabilization of fade-out effects such that infection persist; migration from a neighboring setting generating infection bounce back in locations where infection has been well controlled; and lower PPVs for transmission elimination, proceeding treatment achieving a target prevalence of infection threshold is likely even if all clusters are being treated when migration between locations is occurring. The key result is that migration from an untreated external source (UES) is capable of causing bounce back even in areas with effective MDA coverage to get transmission close to the breakpoint once its rate equals or exceeds the death rate of eggs or larvae in the infectious reservoir. For hookworm, this is typically around 26 infected individuals per year. As a consequence of the result above, if the rate of infected migration exceeds or equals the defined threshold, the PPV for elimination may be lowered even if all clusters are treated.

Finally, mathematical modellers interested in public health will hopefully benefit from open access to simulation models, such as our public code, [helmpy](https://sites.google.com/view/helmpy), in order to examine the effects of infected human migration on the effectiveness of their control policies for STH and other helminths, though some simpler approximate descriptions exist under certain assumptions [35]. In future work, we plan also to extend the [helmpy](https://sites.google.com/view/helmpy) package to include features for likelihood-based inference for parameter estimation from epidemiological data in given settings.

The models suggest the existence of an important critical scale (i.e., order of magnitude) for the migration rate (up to the population number caveat discussed in Sec. 3.1) corresponding to the death rate of eggs or larvae in the case of STH infections in the infectious reservoir exists, above which, simulations exhibit various behaviours. These include stabilization of fade-out effects such that infection persist; migration from an neighboring setting generating infection bounce back in locations where infection has been well controlled; and lower PPVs for transmission elimination, proceeding treatment achieving a target prevalence of infection threshold is likely even if all clusters are being treated when migration between locations is occurring. The challenges for future work on migration impact on the likelihood of achieving transmission breakpoints, lie in good measurement of both the heterogeneity of MDA coverage over time in adjacent health implementation units and migration patterns between such units.

## Data Availability

NA

## Acknowledgements

RJH, MW, JET and RMA gratefully thank the Bill and Melinda Gates Foundation for research grant support via the DeWorm3 (OPP1129535) award to the Natural History Museum in London (http://www.gatesfoundation.org/). The authors would also like to thank Emily McNaughton for project management and helpful comments on the manuscript. The views, opinions, assumptions or any other information set out in this article are solely those of the authors. All authors acknowledge joint Centre funding from the UK Medical Research Council and Department for International Development (MR/R015600/1).

## Appendix A. Derivation and solution of the master equation

The temporally inhomogenous process of an individual’s worm uptake and death is obtained from the Markovian approximation to a summing over the possible histories in the following way

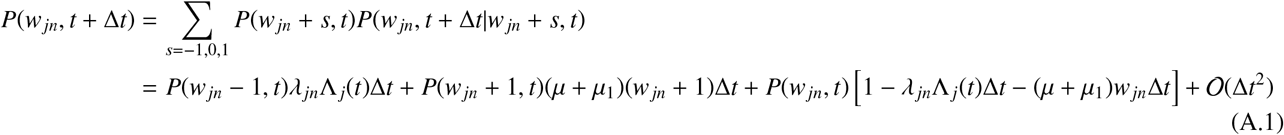

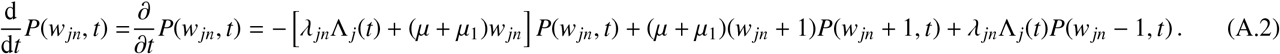

Which matches Eq. (15) in the main text. We may also rewrite Eq. (A.2) using the Probability Generating Function 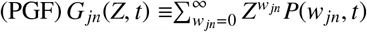 such that

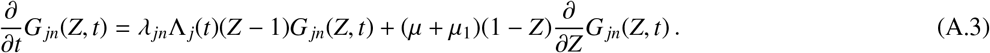

By method of characteristics,^5^ the solution to Eq. (15) is

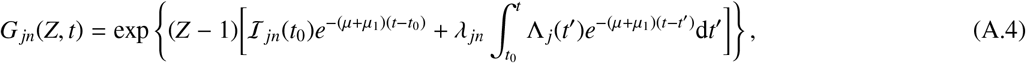

where the initial intensity ℐ_*jn*_(*t*_0_) is to be set for each individual. One immediately recognises Eq. (A.4) as the PGF of an inhomogeneous Poisson walker with intensity

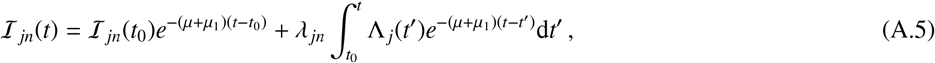

which matches Eq. (16) in the main text.

For an ensemble of *N*_*j*_ independent walkers (hence individuals) in a given age bin we may obtain the PGF of the sum of their collective worm burdens *G*_*j*_(*Z, t*) by multiplication such that

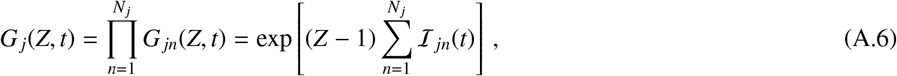

where the corresponding probability mass function is therefore also that of a Poisson distribution

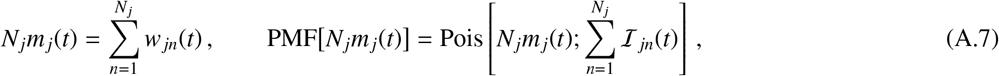

where we have also used the definition of the mean worm burden in the *j*-th age bin given by Eq. (4) in the main text. Note that in the case where Λ _*j*_(*t*) is roughly constant for all time, the timescale for the distribution to achieve stationarity is Δ*t*_stat_ 1/(*µ* + *µ*_1_).

## Appendix B. Approximate ensemble mean and variance of the mean worm burden

The approximate mean and variance of Λ _*j*_(*t*) may be computed by inserting Eqs. (22) and (23) into Eq. (20) and its square, yielding the following equations

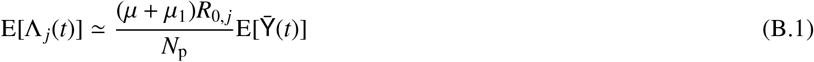

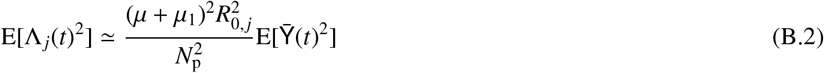

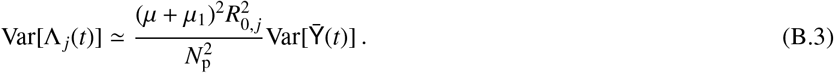

Using Eq. (17) in the main text and Eq. (B.1), we may therefore compute the ensemble mean of *m*_*j*_(*t*)

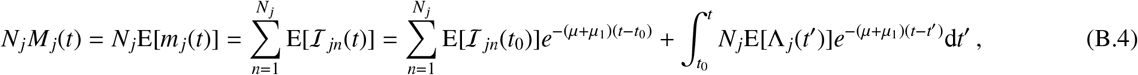

which, by identifying 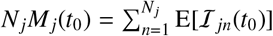, matches the solution to Eq. (12) in the rapid equilibriation limit dE(Λ _*j*_)/d*t* → 0 and hence verifies the consistency of our approximative results in this section so far.

The ensemble variance of *m*_*j*_(*t*) may also be deduced as

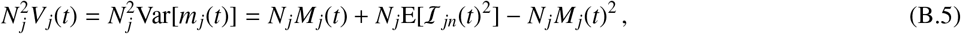

from the properties of independent samples.^6^ By making use of Eq. (1) and assuming the relative independence of λ _*jn*_ and Λ(*t*) once again, we have

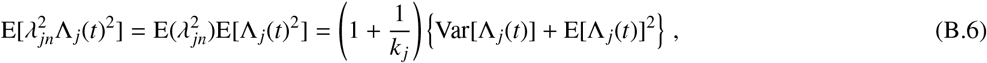

and hence one may obtain E[I _*jn*_(*t*)^2^], which is given by^7^

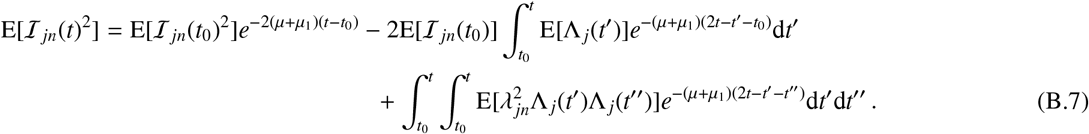

If one assumes temporal stationarity such that E[Λ _*j*_(*t*)] = E(Λ _*j*_) and 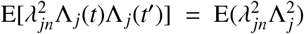, both time integrations in Eq. (B.7) are trivial and one obtains

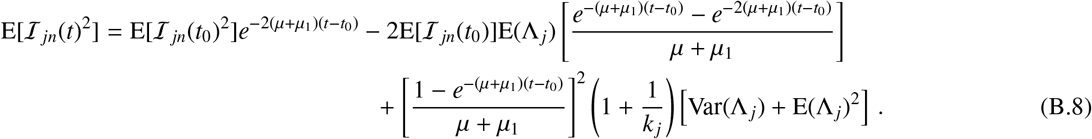

## Appendix C. The van Kampen expansion of the master equation

We may re-derive Eq. (15) in terms of the sum over all worms carried by *N*_*j*_ individuals in the *j*-th age bin 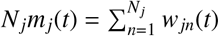, taking a single step at each point in time, such that

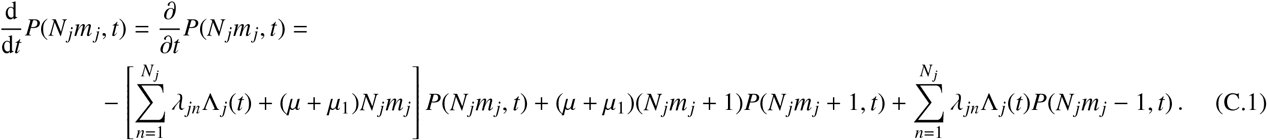

Though this equation would almost-certainly represent a less efficient simulation than that of Eq. (15), it may nonetheless consistent results for a sufficiently short choice of numerical timestep.

Let us now rewrite the local value of *N*_*j*_*m*_*j*_ (which is regarded as constant in time — or a ‘state variable’ — by the master equation) in separate components which depend differently on the system size *N*_*j*_, to obtain

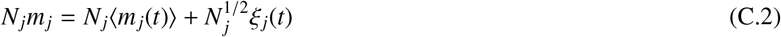

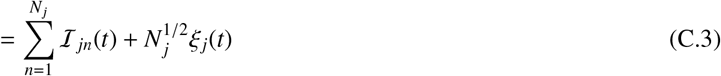

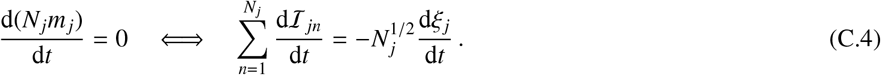

Since the mean of Eq. (27) is an averaged quantity it does not fluctuate as a random variable, hence we may now rewrite the probability distribution in terms of the other remaining random variable which now characterises the fluctuations of the system around the temporal average

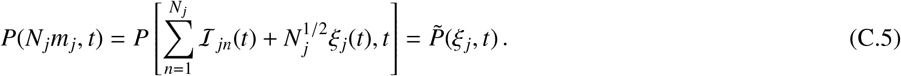

The total time-derivatives of both distributions are hence related, yielding

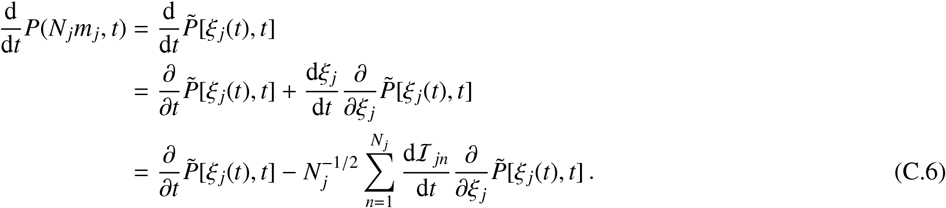

Notice that Eq. (27) implies steps of *N*_*j*_*m*_*j*_ ± 1 translate into fluctuations of 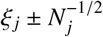 in the argument of 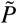. Using this mapping as well as Eq. (27), one finds that Eq. (C.1) may be rewritten as

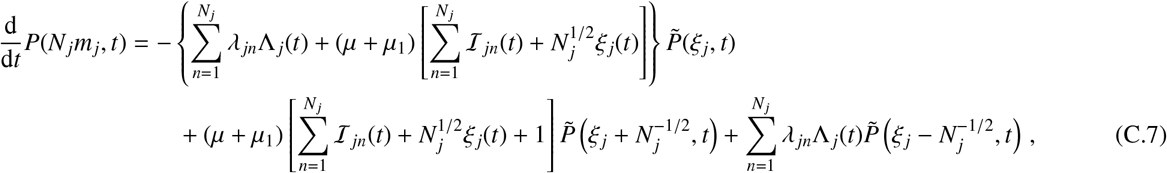

where the following van Kampen [38] (or Taylor) expansion of the distributions on the right hand side can be made which is controlled by powers of the system size *N*_*j*_

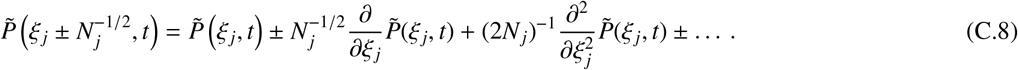

By equating Eq. (C.6) and Eq. (C.7) with the expansion above, we may match terms of the same system size order to obtain constraint equations. At 𝒪(*N_j_*) there is a cancellation of terms, followed by 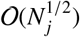 where the mean-field results of Eqs. (12), (13) and (14) are consistently reproduced after *a posteriori* marginalisation over λ _*jn*_ values. Lastly, at 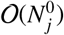, one obtains a linear noise approximation for the fluctuations in the form of a Fokker-Planck equation

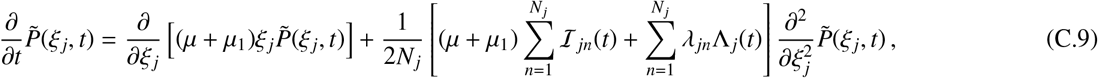

which also corresponds to the following Langevin equation for the individual realisations

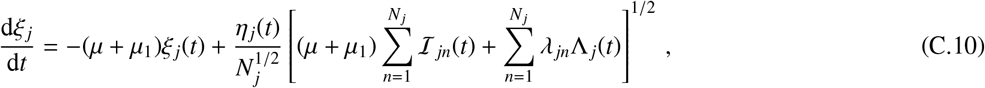

where *η* _*j*_(*t*) is a zero-mean unit-amplitude Gaussian random variable with the *temporal* averages of ⟨*η* _*j*_(*t*) ⟩= 0, ⟩*η* _*j*_(*t*)*η* _*j*_(*t*,) ⟩= δ(*t* − *t′*).

## Footnotes

1 More generally, to include a mean of *X*, this distribution is modified to Gamma(λ _*jn*_; *k* _*j*_, *k* _*j*_/*X*).

2 Note, e.g., that a compound Poisson process with logseries-distributed jumps also produces a negative binomial distribution. For a recent example which applys this process in the context of disease transmission, see: https://nbviewer.jupyter.org/github/umbralcalc/covid-simple/blob/master/covid-simple.ipynb.

3 The code is also accompanied by an interactive notebook of examples matching the calculations made in this paper which may be found in the

4 As was also shown in Ref. [35], when the reservoir timescale *µ*_2_ is not as fast, Eq. (34) should receive a non-Markovian modification ∝ 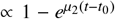 to account for the slower decay of pulses in the reservoir.

5 One solves first-order PDEs of the form in Eq. (A.3) by method of characteristics, i.e., choosing a curve *Z*(*t*) such that 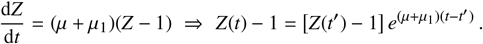 Given the fact that along the curve *Z*(*t*) one also obtains an effective ODE of the form 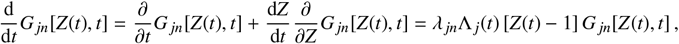 whose solution is given by Eq. (A.4) when one replaces *Z*(*t*,) − 1 with the inverted form of the equation above.

6 Note that one finds 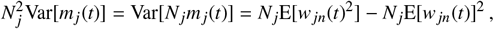 so Eq. (B.5) is consistent with the number of random degrees of freedom present.

7 Note that to obtain the relation 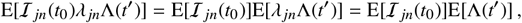 in the second term of Eq. (B.7) we have assumed that the initial intensities I _*jn*_(*t*_0_) are chosen independently of an individual’s uptake rate λ _*jn*_.

